# A Cross Sectional Study of Atrial Fibrillation Prevalence and Associated Cardiac Arrhythmias in Stroke Patients: Insights from Commercial Data Archives

**DOI:** 10.1101/2024.04.21.24306150

**Authors:** Kuan-Meng Soo, Te-Sheng Liew, Paul Heath

## Abstract

The link between atrial fibrillation (AF) and stroke has gained attention due to AF’s significant impact on stroke risk. Detecting AF, especially paroxysmal AF, presents challenges, highlighting the necessity for continuous Holter monitoring. In our investigation, we utilized commercial data archives to examine AF prevalence and associated cardiac arrhythmias among stroke patients and controls. We also analyzed various factors, such as age, gender, symptoms, test duration, and duration post-stroke, to identify predictors of arrhythmias and enhance paroxysmal detection rates. We included 98 stroke patients and 98 controls for comparing AF prevalence within 72 hours post-stroke and analyzed 779 stroke patients within 4 days to 10 weeks post-stroke. Our findings revealed a higher AF detection rate in stroke patients compared to controls, with AF being the most prevalent arrhythmia observed. Older age and symptom presence were associated with significant findings, particularly AF. Temporal analysis showed a negative correlation between AF and complete heart block occurrence over time, while ventricular tachycardia exhibited a positive correlation. Paroxysmal fibrillation detection rates were highest within the initial month, with the optimal duration of Holter monitoring being 10 days, underscoring the importance of extended monitoring for detecting paroxysmal atrial fibrillation. Detecting paroxysmal atrial fibrillation could influence stroke management. Furthermore, our study suggests the potential utility of commercial data in yielding comparable outcomes to research data.

## INTRODUCTION

There is a growing awareness of the association between atrial fibrillation (AF) and stroke. AF, characterized by irregular atrial contractions, leads to ineffective blood pumping, causing blood stasis and increasing the risk of clot formation. Clots that travel to the brain can result in ischemic strokes, which are common and often devastating, with 70-80% of patients experiencing severe outcomes (Gladstone et al., 2014; Pistoia et al., 2016).

The presence of atrial fibrillation (AF) may indicate the need for anticoagulants to prevent strokes (Tanislav & Kostev, 2019). However, paroxysmal AF, particularly, may not be evident before a stroke, except in cases where patients have implanted devices (Baturova, Lindgren, Shubik, et al., 2014). Paroxysmal AF is suspected as an underlying cause of stroke, with similar or potentially higher stroke rates compared to sustained AF (Fujii et al., 2013; Giralt-Steinhauer et al., 2015). Continuous Holter monitoring is necessary to capture these episodes, and commercial Holter archives can support research efforts.

We utilize commercial data archives to evaluate stroke and associated cardiac arrhythmia findings for a comprehensive understanding of AF prevalence among stroke patients. Initially, we compare AF prevalence within 72 hours post-stroke with age and gender-matched controls, assuming AF may have developed prior to stroke. Secondly, we stratify stroke patients based on age, gender, symptoms, test duration, and duration post-stroke to identify factors closely associated with arrhythmia and improve paroxysmal AF detection rates, minimizing resource consumption during patient monitoring.

Lastly, we investigate the detection rate of arrhythmias in relation to testing duration and time post-stroke. Given the lack of consensus on the optimal monitoring duration for AF detection and limited data addressing this question, our study aims to provide insights into this area.

## METHOD

### Study design and setting

This study utilized data from the Cardioscan Pye Ltd database. The data spanned from January 2021 to December 2023, and upon examination, it met the criteria for inclusion in the study, encompassing participants from Malaysia and the United States.

### Ethical considerations

The study adhered to the principles outlined in the Declaration of Helsinki. Patient privacy and confidentiality were safeguarded through the deidentification of patient records, and no interventions were administered to eliminate ethical concerns.

### Participants

The participants consisted of individuals with a confirmed history of stroke, including those who had experienced a stroke within the past 0-3 days and within the past 10 weeks. Data from stroke patients and controls were collected from individuals who underwent three-channel Holter ECG monitoring (myPatch-sl) at various hospitals or clinics. The results were then sent to Cardioscan Pye Ltd for analysis. Controls were chosen through stratified random sampling from the database to match the age and gender distributions of the stroke patients.

### Variables

The study comprised three parts. In the first part, the detection rate of significant Holter findings in stroke patients was compared with controls. The second part involved comparing age, gender, symptoms, and specific symptoms (palpitations, syncope, and dizziness) between groups with and without significant Holter findings. The third part compared the detection rate of significant Holter findings among stroke patients at different weeks post-stroke and with different durations of Holter tests.

Information was sourced from the Cardioscan Pye Ltd database, which aggregated data including significant Holter findings, age, gender, stroke symptoms, days since stroke occurrence, and duration of Holter tests for stroke patients. ECG results were evaluated using Cardioscan 12 Satellite software after excluding recordings with artifacts. Trained cardiac physiologists at Cardioscan Pye Ltd classified post-stroke patients into categories based on the presence or absence of significant findings. This methodology aims to reduce the potential for bias stemming from assessors lacking adequate expertise.

### Statistical analyses

Normality was assessed using the Shapiro-Wilk test in JASP 0.18.2, with a significance level set at p < 0.05 indicating non-normal distribution. Age data were presented as median and interquartile range. Mann-Whitney tests, conducted using JASP 0.18.2, were employed to compare age differences between groups with and without significant findings. Gender, symptom, and significant finding variables were expressed as counts and percentages. Differences in male percentage, symptom presence, and significant findings presence between groups were evaluated using chi-square tests. The chi-square test was performed using an online calculator (Social Science Statistics, 2024), with a significance level set at p < 0.05. Multivariate logistic regression analysis was conducted using JASP 0.18.2. Spearman correlation analyses were carried out to examine the correlation between duration post-stroke and the detection rate of significant findings, as well as between test duration and the detection rate of significant findings.

## RESULTS

### Records selection process

In this study, the process of selecting records comprised enrolling a total of 98 patients and 98 controls for the first and second parts of the study, respectively. The median age and gender distribution are presented in Table 1. For the third part of the study, 779 patients were included after eliminating records that did not meet the inclusion criteria or lacked relevant information, as illustrated in Figure 1. The median age was 67 years, with an interquartile range spanning 20 years. Among these participants, 438 were male and 347 were female.

**Table 1.** comparison between stroke and control group

|  | Stroke group | Control group | p-value |
| --- | --- | --- | --- |
| N | 98 | 98 | - |
| Gender | 41 (41.8) | 41 (41.8) | 1.000 |
| Age | 69 (26) | 62 (28) | 0.556 |
| AF | 20 (20.4) | 1 (1) | $<0.05$ |
| PA | 0 (0) * | 3 (3) | - |
| PF | 3 (3.1) | 3 (3) | 0.990 |
| VE | 0 (0) * | 1 (1) | - |
| VT | 2 (2) | 4 (4) | 0.414 |
| SD | 0 (0) * | 1 (1) | - |
| SV | 1 (1) | 2 (2) | 0.567 |

\* Chi-square analysis was not feasible due to the absence of significant findings in the dataset.
AF (Atrial Fibrillation, with heart rate $> 110$ rpm), PA (pauses, greater than 3 seconds), PF (Paroxysmal/Sustained Atrial Fibrillation), VE (Very Frequent Ventricular Ectopics, VE burden $>10\%$ ), VT (Ventricular Tachycardia, 6 beats or more), SD (2:1 Second Degree AV Block or High Degree AV Block), SV (Sustained Supraventricular Tachycardia)

**Figure 1.**
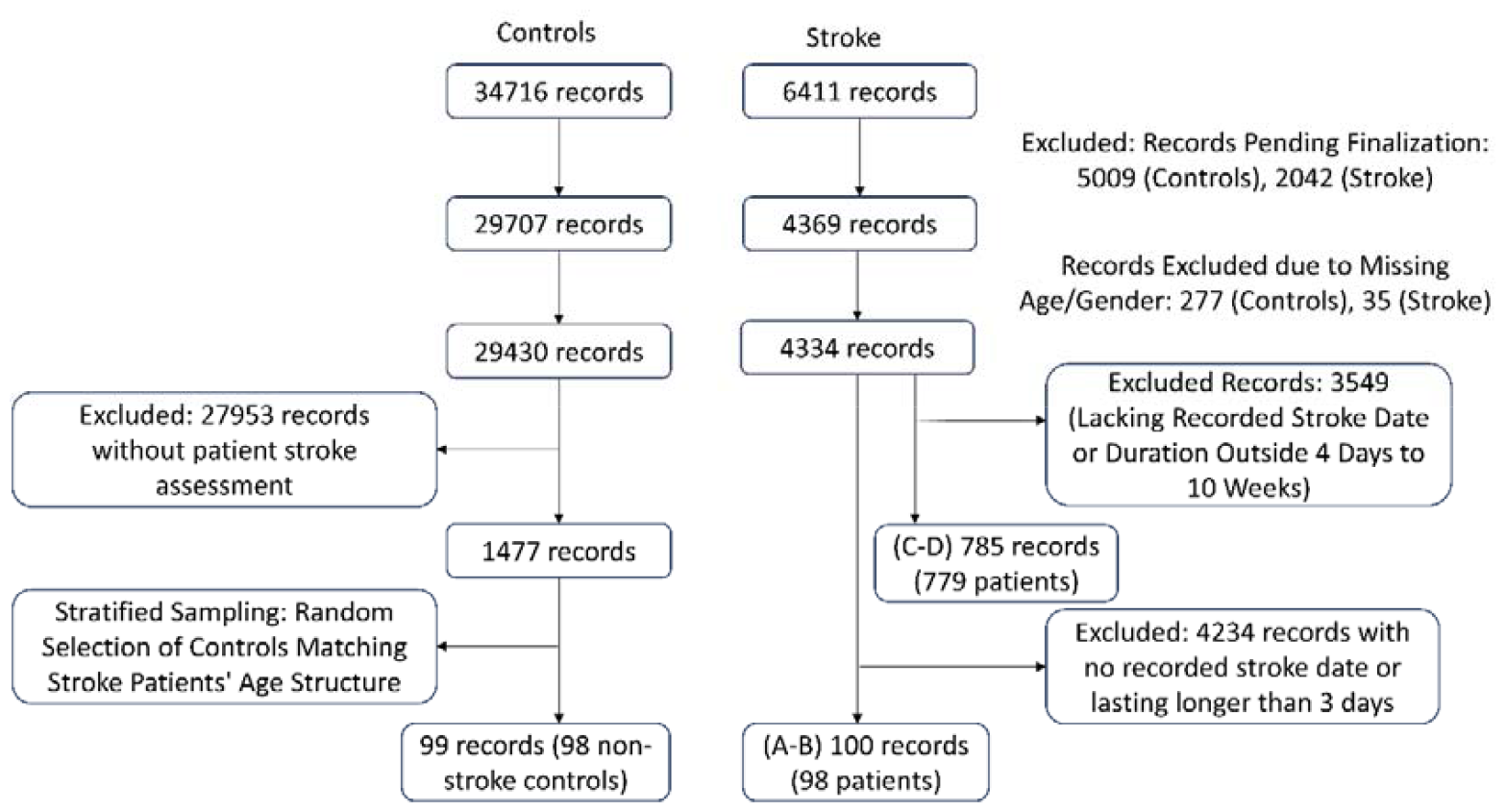
Records selection flow chart

### Comparison between stroke and control group

Table 1 presents the results of significant findings in both the stroke and control groups. It was observed that the detection rate of atrial fibrillation (AF) was higher in the stroke group compared to the control group (p-value < 0.05), while the remaining significant findings did not show a difference between the two groups. The stroke group exhibited atrial fibrillation as the most prevalent arrhythmia, followed by paroxysmal atrial fibrillation and ventricular tachycardia in descending order of frequency.

### Demographic and clinical correlates: age, gender, and symptoms in various significant findings

Table 2 illustrates the comparisons between the presence and absence of various significant findings. Patients with significant findings, particularly AF, were found to be older, have a higher percentage of males, and a higher incidence of symptoms. Most comparisons did not yield significant differences, likely due to the low sample size.

**Table 2.** Comparisons between presence and absence of various significant findings

|  | N | Age, median (IQR)*** | Gender (%) | Symptoms (%) | Palpitations (%) | Syncope (%) | Dizziness (%) |
| --- | --- | --- | --- | --- | --- | --- | --- |
| Total significant finding | 26 | 75 (10)<sup>a</sup> | 11 (42.3)<sup>a</sup> | 4 (15.4) | 3 (11.5) | 0 (0)<sup>**</sup> | 1 (3.8) |
| without significant finding | 72 | 60 (29) | 30 (41.7) | 25 (34.7) | 14 (19.4) | 5 (6.9) | 3 (4.2) |
| With AF or PF | 23 | 74 (10)<sup>a</sup> | 11 (47.8)<sup>a</sup> | 2 (8.7) | 1 (4.3) | 0 (0) | 1 (4.3) |
| Without AF or PF | 75 | 62 (30) | 30 (40) | 27 (36) | 16 (21.3) | 5 (6.7) | 3 (4) |
| with AF | 20 | 75 (11)<sup>a</sup> | 9 (45) | 1 (5)<sup>a</sup> | 0 (0)<sup>**</sup> | 0 (0)<sup>**</sup> | 1 (5) |
| without AF | 78 | 63 (29) | 32 (41) | 28 (35.9) | 17 (21.8) | 5 (6.4) | 3 (3.8) |
| with PF | 3 | 70 (3) | 2 (66.7) | 1 (33.3) | 1 (33.3) | 0 (0)<sup>**</sup> | 0 (0)<sup>**</sup> |
| without PF | 95 | 68 (27) | 39 (41.1) | 28 (29.5) | 16 (16.8) | 5 (5.3) | 4 (4.2) |
| with SV | 1 | 75 (0)<sup>*</sup> | 0 (0)<sup>*</sup> | 0 (0)<sup>**</sup> | 0 (0)<sup>**</sup> | 0 (0)<sup>**</sup> | 0 (0)<sup>**</sup> |
| without SV | 97 | 69 (26) | 41 (42.3) | 29 (29.9) | 17 (17.5) | 5 (5.2) | 4 (4.1) |
| with VT | 2 | 73 (7) | 0 (0)<sup>*</sup> | 2 (100)<sup>**</sup> | 2 (100)<sup>**</sup> | 0 (0)<sup>**</sup> | 0 (0)<sup>**</sup> |
| without VT | 96 | 69 (27) | 41 (42.7) | 27 (28.1) | 15 (15.6) | 5 (5.2) | 4 (4.2) |

AF (Atrial Fibrillation, with heart rate > 110rpm), PF (Paroxysmal/Sustained Atrial Fibrillation), SV (Sustained Supraventricular Tachycardia), VT (Ventricular Tachycardia, 6 beats or more)
<sup>a</sup> represent significant different from without the significant finding, with p-value < 0.05
<sup>\*</sup> chi-square not able to be carried out due to 0 female or 0 male in these significant findings.
<sup>\*\*</sup> chi-square not able to be carried out due to 0 symptoms or 0 without symptoms in these significant findings.
<sup>\*\*\*</sup> the confidence interval for age is available in Appendix IV of the supplementary file.

Following multivariate logistic regression analysis (Table 3), age remained a significant predictor for total significant findings, AF or paroxysmal Atrial fibrillation (PF), and AF alone. Gender was not significant for predicting significant findings, while symptoms were significant predictors for AF or PF and AF alone.

**Table 3.** Multivariate logistic regression model.

| Significant findings | Age |  | Gender |  | Symptom |  |
| --- | --- | --- | --- | --- | --- | --- |
|  | OR (95% CI) | Standardized | OR (95% CI) | Standardized | OR (95% CI) | Standardized |
| Total significant finding | 1.068 (0.027 – 0.104)<sup>*</sup> | 3.331 | 0.818 (-1.189 – 0.786) | -0.399 | 0.294 (-2.473 – 0.027) | -1.918 |
| AF or PF | 1.064 (0.022 – 0.103)<sup>*</sup> | 3.051 | 1.113 (-0.913 – 1.127) | 0.206 | 0.148 (-3.489 – -0.327)<sup>*</sup> | -2.365 |
| AF | 1.064 (0.020 – 0.105)<sup>*</sup> | 2.869 | 0.933 (-1.136 – 0.997) | -0.128 | 0.079 (-4.644 – -0.433)<sup>*</sup> | -2.363 |
| PF | 1.038 (-0.053 – 0.128) | 0.812 | 2.597 (-1.544 – 3.453) | 0.749 | 1.614 (-2.033 – 2.991) | 0.374 |
| SV | 1.070 (-0.130 – 0.265) | 0.670 | 6.8*10<sup>-9</sup> (-1.3*10<sup>4</sup> – 1.3*10<sup>4</sup>) | -0.003 | 1.1*10<sup>-8</sup> (-1.6*10<sup>4</sup> – 1.6*10<sup>4</sup>) | -0.002 |
| VT | 1.073 (-0.066 – 0.207) | 1.014 | 0.000 (-1.2*10<sup>4</sup> – 1.2*10<sup>4</sup>) | -0.003 | 3.3*10<sup>8</sup> (-9.8*10<sup>3</sup> – 9.8*10<sup>3</sup>) | 0.004 |

AF (Atrial Fibrillation, with heart rate > 110rpm), PF (Paroxysmal/Sustained Atrial Fibrillation), SV (Sustained Supraventricular Tachycardia), VT (Ventricular Tachycardia, 6 beats or more)
<sup>\*</sup> represents p- value < 0.05

Figure 2 depicts the temporal distribution of individuals with significant ECG findings following a stroke. Spearman’s correlation analysis revealed a negative correlation between the occurrence of AF and complete heart block (CB) with the number of weeks post-stroke, resulting in correlation coefficients of -0.769 and -0.683, respectively. Specifically, AF decreased from a detection rate of 11.5% in the first week to between 0-2% starting from the third week post-stroke. Conversely, CB decreased from 0.5-0.6% in the first and second weeks to a 0% detection rate in the third week post-stroke. In contrast, ventricular tachycardia (VT) exhibited a positive correlation, with a correlation coefficient of 0.697. VT increased from 2.8% in the first week to 9.5% and 8.3% in the ninth and tenth weeks post-stroke. The remaining ECG findings did not demonstrate significant correlations with the number of weeks post-stroke.

**Figure 2.**
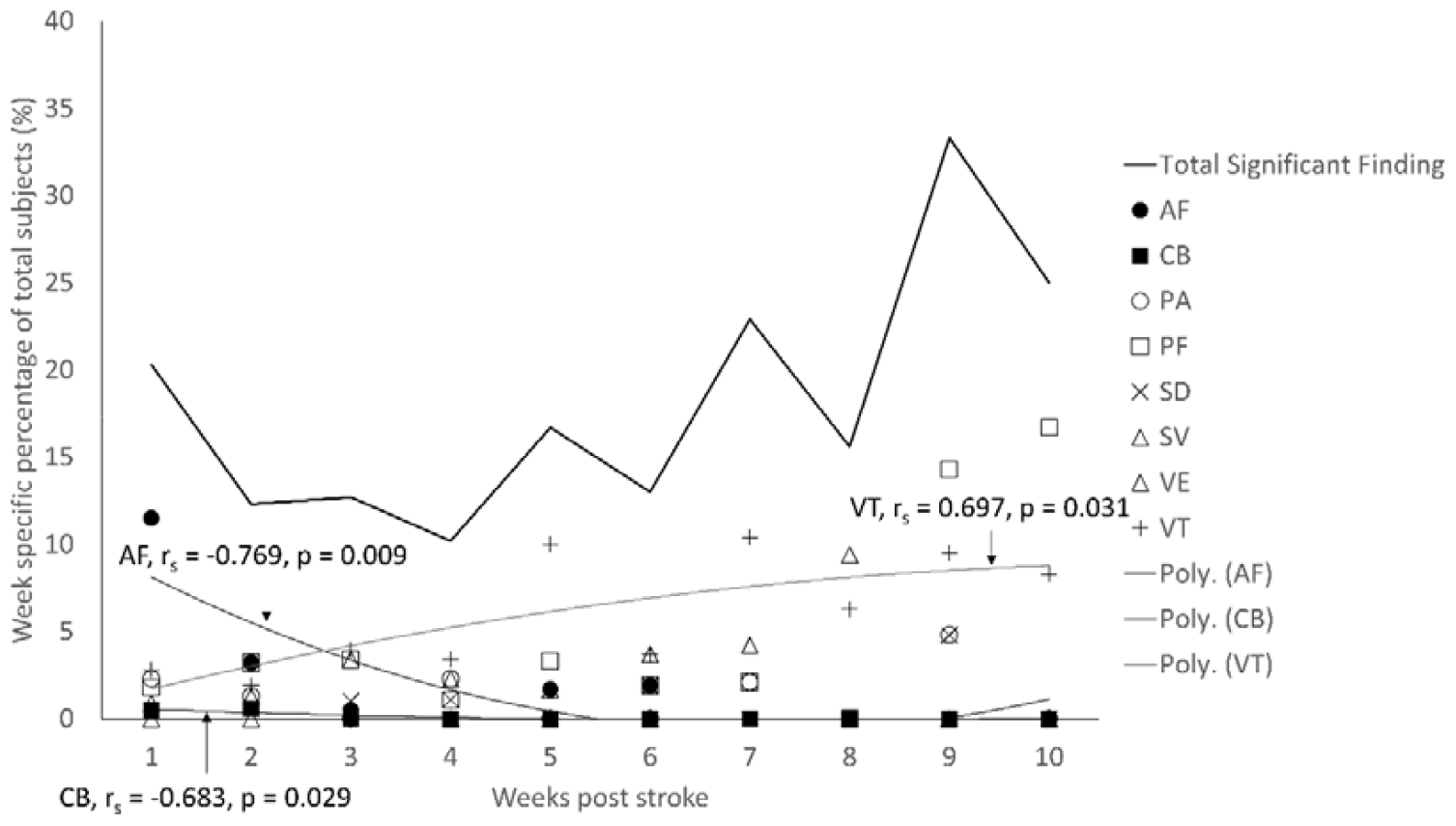
Week specific distribution of individuals with significant ECG findings post stroke. AF (Atrial Fibrillation, with heart rate > 110rpm), CB (Complete Heart Block), PA (Pauses, greater than 3 seconds), PF (Paroxysmal/Sustained Atrial Fibrillation), SD (2:1 Second Degree AV Block or High Degree AV Block), SV (Sustained Supraventricular Tachycardia), VE (Very Frequent Ventricular Ectopics, VE burden >10%) and VT (Ventricular Tachycardia, 6 beats or more). The data that is used to plot this figure is available in Appendix II of the supplementary file.

Figure 3 illustrates the detection rate of AF, PF, and other significant findings (CB, PA, PF, SD, SV, VE, VT) across different durations of Holter monitoring and duration since stroke onset, while excluding significant findings occurring within the initial 3 days following the stroke onset. AF was not detected until more than 3 days after stroke onset in 35% of cases (11 out of 31), while PF was not detected until more than 3 days after stroke onset in 88% of cases (21 out of 24). The highest detection rate of atrial fibrillation (AF) was observed in Holter monitoring tests lasting for 2 days within the first month following stroke onset. Similarly, the highest detection rate of paroxysmal fibrillation (PF) was noted in Holter monitoring tests lasting for 10 days within the initial month after stroke onset.

**Figure 3.**
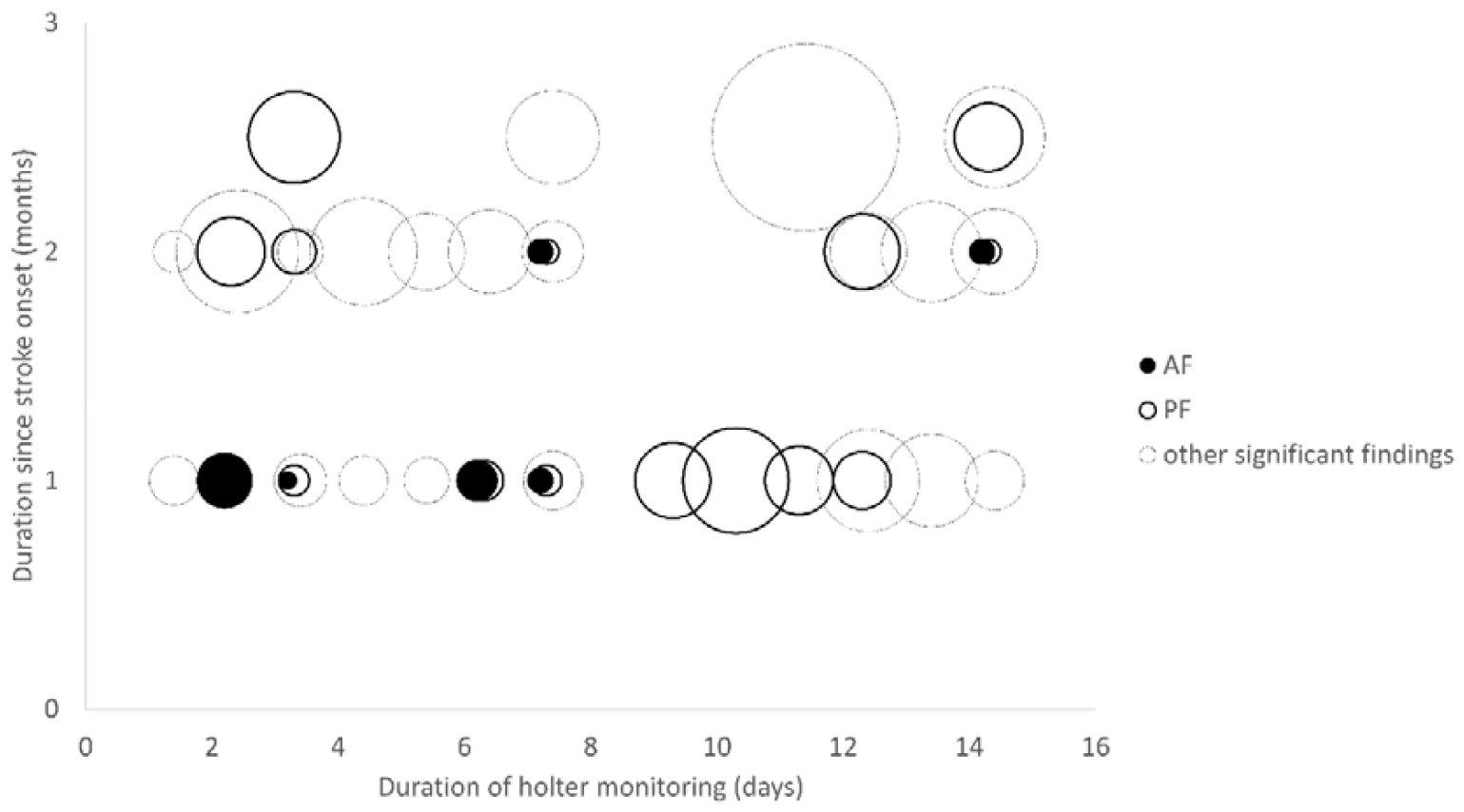
Association Between Time Post-Stroke and Holter Test Duration with Arrhythmia Detection Rate: Bubble Plot Analysis. AF (Atrial Fibrillation, with heart rate > 110rpm), CB (Complete Heart Block), PA (Pauses, greater than 3 seconds), PF (Paroxysmal/Sustained Atrial Fibrillation), SD (2:1 Second Degree AV Block or High Degree AV Block), SV (Sustained Supraventricular Tachycardia), VE (Very Frequent Ventricular Ectopics, VE burden >10%) and VT (Ventricular Tachycardia, 6 beats or more). The data that is used to plot this figure is available in Appendix III of the supplementary file.

Table 4 presents the results of multiple linear regression. No significant regression was found between the variables. However, it was observed that the duration since stroke onset had a greater effect on AF detection compared to the duration of Holter monitoring (−0.287 vs -0.196), while for PF, the duration of Holter monitoring had a greater effect on the detection rate compared to the duration since stroke onset (0.128 vs -0.088). For other significant findings, both the duration since stroke onset and the duration of Holter testing had similar effects on the detection rate (0.229 vs 0.289).

**Table 4.** Multiple linear regression

| Significant findings | Duration of holter monitoring |  | Duration since stroke onset |  |
| --- | --- | --- | --- | --- |
|  | Estimates (95% CI) | Standardized | Estimate | Standardized |
| AF | -0.0008 (-0.0002 – 0.0006) | -0.196 | -0.008 (-0.018 – 0.002) | -0.287 |
| PF | 0.003 (-0.005 – 0.010) | 0.128 | -0.012 (-0.061 – 0.037) | -0.088 |
| Other significant findings | 0.011 (-0.005 – 0.026) | 0.229 | 0.092 (-0.016 – 0.200) | 0.289 |

## DISCUSSIONS

### Detection rate of atrial fibrillation in stroke patients versus controls

The detection rate of atrial fibrillation (AF) is higher in stroke patients compared to controls, consistent with findings from previous studies (Baturova, Lindgren, Carlson, et al., 2014) (Baturova, Lindgren, Shubik, et al., 2014) (Tanislav & Kostev, 2019).

### Factors impacting the prevalence of atrial fibrillation among stroke patients

Our research observed a slightly higher prevalence of atrial fibrillation (AF) and paroxysmal fibrillation (PF) among post-stroke patients (0.23 ± 0.05) compared to earlier studies (0.16 ± 0.02) (refer to appendix III for the detail of this prevalence) (Wohlfahrt et al., 2014)(Fujii et al., 2013)(Sutamnartpong et al., 2014)(Yoshioka et al., 2015)(Giralt-Steinhauer et al., 2015)(Kamel et al., 2009) (Amaya Pascasio et al., 2022). This difference might be attributed to previous studies relying on registry data without comprehensive ECG documentation, potentially leading to an underestimation of AF and PF rates (Baturova, Lindgren, Shubik, et al., 2014).

Additionally, stroke patients with AF or PF tended to be older and exhibited more symptoms. Aging induces structural changes in the heart, such as increased fibrosis and scarring, disrupting normal electrical conduction and contributing to arrhythmias (Kistler et al., 2004). Interestingly, while AF prevalence increases with age in both stroke patients and controls, prior research has indicated that the hazard ratio of AF diminishes with age. This suggests that the gap in AF detection rates between stroke patients and controls lessens with age, making it uncertain whether AF is indicative of recurrent stroke or simply a consequence of aging. In essence, AF seems to be more readily identified as a sign of stroke in younger individuals compared to older ones, as the disparity in detection rates between stroke patients and controls diminishes with age (Kamel et al., 2009).

While symptoms, notably palpitations, frequently indicate underlying cardiac rhythm disturbances, our study did not identify a significant association with specific symptoms such as palpitations, despite demonstrating an association with symptoms in general. However, prior studies have shown a strong correlation between palpitations and arrhythmias, particularly supraventricular (SV) arrhythmias (Helton, 2015).

### Temporal patterns of cardiac arrhythmias following stroke

The peaks of atrial fibrillation (AF) and complete heart block (CB) were noticeable within the first two days following a stroke, which aligns with research indicating significant cardiac adverse events typically occurring between days 2 and 3 post-stroke. This emphasizes the crucial role of continuous monitoring, including Holter monitoring beyond the initial 24 hours (Kallmünzer et al., 2012).

The decrease in AF detection over time among patients could be attributed to various treatments such as radiofrequency ablation (RFA), antiarrhythmic drugs, or oral anticoagulants, which are known to effectively manage AF (Tanislav & Kostev, 2019; Waks & Zimetbaum, 2017; Yoshida et al., 2010). Antiarrhythmic drugs can suppress AF, facilitating long-term maintenance of sinus rhythm, while RFA can reduce the dominant frequency of AF and alleviate complete heart block (Boulos et al., 2004). Moreover, a substantial proportion of stroke patients with AF received oral anticoagulant therapy, though not universally (Tanislav & Kostev, 2019).

The declining rates of AF and CB detection might also be influenced by fewer studies conducted over extended post-stroke periods, potentially limiting opportunities for identification. However, ventricular tachycardia tends to increase post-stroke over time, possibly due to the side effects of antiplatelet therapy or stroke-induced autonomic dysregulation, leading to alterations in heart rate. Additionally, the stress response and sympathetic activation associated with stroke could further contribute to tachycardia (Matsuzono et al., 2023).

The detection of paroxysmal fibrillation (PF) was found to increase after 3 days post-stroke, with the majority of cases being detected beyond this timeframe (88%, 21/24). PF also showed a higher detection rate during Holter testing with longer durations. Most PF cases remained undetected until after 3 days post-stroke onset, indicating delayed PF manifestation and highlighting the importance of extended monitoring for PF detection. These findings are consistent with a study by Kamel et al., which monitored patients for up to 90 days post-stroke, revealing that most PF cases were detected after 3 days post-stroke onset (Kamel et al., 2009). However, our results differ from those of Sutamnartpong et al., where most PF cases occurred within 24 hours, although the duration of the study relative to stroke onset was unspecified (Sutamnartpong et al., 2014).

Despite conducting an ad hoc association study between PF and symptoms (Table 5), we found no significant difference in symptom presence between individuals with and without AF or PF. This suggests that PF detection may be challenging, lacking distinguishing symptoms compared to individuals without PF. Gladstone’s study further supports the efficacy of prolonged monitoring in increasing AF detection rates, demonstrating twice the detection rate after 4 weeks compared to 1-week monitoring (14.8% vs. 7.4%). This increased detection could result in more patients receiving anticoagulant therapy, ultimately reducing the risk of secondary stroke (Gladstone et al., 2014). These findings are consistent with recommendations in stroke management guidelines, advocating for prolonged monitoring of approximately 30 days with Holter monitoring within 6 months post-stroke for strokes without a known cause (Edwards et al., 2016).

**Table 5.** Comparisons between presence and absence of various significant findings, after 3 days since stroke onset until 10 weeks.

|  | N | Symptoms (%) | Palpitations (%) | Syncope (%) | Dizziness (%) |
| --- | --- | --- | --- | --- | --- |
| With AF or PF | 32 | 5 (15.6) | 4 (12.5) | 1 (3.1) | 0 (0) ** |
| Without AF or PF | 747 | 113 (15.1) | 70 (9.4) | 11 (1.5) | 17 (2.3) |
| with AF | 11 | 0 (0) ** | 0 (0) ** | 0 (0) ** | 0 (0) ** |
| without AF | 768 | 118 (15.4) | 74 (9.6) | 12 (1.6) | 17 (2.2) |
| with PF | 21 | 5 (23.8) | 4 (19) | 1 (4.8) | 0 (0) ** |
| without PF | 758 | 113 (14.9) | 70 (9.2) | 11 (1.5) | 17 (2.2) |

\*\* chi-square not able to be carried out due to 0 symptoms or 0 without symptoms in these significant findings.

### Limitations

One limitation of this study pertains to potential variability in symptom reporting by referring physicians, which might result in incomplete data on symptoms and potentially underestimate their prevalence among stroke patients. Additionally, the diversity in treatment and management received by patients following a stroke could influence the study outcomes, although this aspect was not explored in this study. Hence, readers should consider these factors when evaluating the generalizability of the study findings.

A notable portion of the data was excluded due to missing stroke dates, leading to a reduced sample size for the study. Furthermore, the data lacked detailed information on the types of strokes, which could include hemorrhagic strokes less commonly associated with AF. This limitation might contribute to some degree of underestimation of AF prevalence among stroke patients. Future research should include stroke type classification to address this issue.

Additionally, the correlation analysis of significant ECG findings’ detection rate post-stroke was restricted to a period up to week 10 due to an insufficient number of studies available for analysis beyond this timeframe.

## CONCLUSION

The incidence of atrial fibrillation (AF) is more pronounced in stroke patients compared to control groups. This elevated occurrence of AF among stroke patients is linked to advanced age and the presence of symptomatic manifestations. While AF detection rates are initially high following a stroke, they tend to diminish over time. Conversely, paroxysmal atrial fibrillation tends to manifest after 3 days post-stroke, with greater detection rates observed through longer-duration Holter monitoring. This underscores the significance of prolonged monitoring for effectively identifying paroxysmal atrial fibrillation. The heightened detection of paroxysmal atrial fibrillation has the potential to influence the management of post-stroke patients and thereby mitigate the risk of recurrent stroke. Additionally, our findings suggest that utilizing commercial data can yield outcomes comparable to those obtained from research data, hinting at its prospective utility for forthcoming studies.

## Data Availability

All data produced in the present work are contained in the manuscript

## Acknowledgement

The authors express their gratitude to Cardioscan Pye Ltd for generously covering the publication fee for this journal. Additionally, they acknowledge Dr. Thungar Visagathilagar for providing valuable comments on the article.

## Funding

Funding for this study was provided by Cardioscan Pye Ltd. The role of the funder was solely in financial support and did not extend to study design, data collection, analysis, interpretation, or manuscript preparation.

## Disclosure of interest

None declared

## Data availability statement

The data can be found in the attached file.

## Supplementary files

Appendix I STROBE guidelines checklist

Appendix II Confidence interval of age in table 2 of result

Appendix III

## Notes

### Competing Interest Statement

The authors have declared no competing interest.

