## Supplemental file for "A Cross Sectional Study of Atrial Fibrillation Prevalence and Associated Cardiac Arrhythmias in Stroke Patients: Insights from Commercial Data Archives"

Appendix I

STROBE guidelines

| **Section/topic** | **Item number** | **Recommendation** | **Page** |
| --- | --- | --- | --- |
| Title and abstract | 1 | Indicate the study’s design with a commonly used term in the title or the abstract  Provide in the abstract an informative and balanced summary of what was done and what was found | The study design, labeled as a "cross-sectional study," was explicitly mentioned in both the title and abstract. |
| Introduction |  |  |  |
| Background/rationale | 2 | Explain the scientific background and rationale for the investigation being reported | The link between atrial fibrillation and stroke is widely recognized. Detecting AF is crucial as it may signal the necessity for anticoagulant therapy to prevent strokes. Utilizing commercial data archives offers a viable approach to assess stroke-related arrhythmia findings. |
| Objectives | 3 | State specific objectives, including any prespecified hypotheses | In this study, commercial data is employed to assess AF prevalence disparities between stroke patients and controls, discern factors linked to arrhythmias in stroke patients, and explore how AF detection rates correlate with test duration and time post-stroke.  Our predetermined hypotheses anticipate a greater AF prevalence among stroke patients versus controls, with AF susceptibility potentially influenced by age, symptoms, and gender. Additionally, we hypothesize that AF detection rates may be affected by duration post-stroke and test duration. |
| Methods |  |  |  |
| Study design | 4 | Present key elements of study design early in the manuscript | The manuscript detailed participants who had experienced a stroke, providing information on their age and gender distribution. Three-channel Holters were employed for data collection. Ethical considerations, study protocols, and statistical analyses were predetermined. |
| Setting | 5 | Describe the setting, locations, and relevant dates, including periods of recruitment, exposure, follow-up, and data collection | Data were collected globally, spanning countries such as Malaysia and the United States. Recruitment, follow-up, and data collection occurred from January 2021 to December 2023, with exposure (stroke) predating the initiation of the study. |
| Participants | 6 | Cohort study - give the eligibility criteria, and the sources and methods of selection of participants; describe methods of follow-up |  |
|  |  | Case-control study - give the eligibility criteria, and the sources and methods of case ascertainment and control selection; give the rationale for the choice of cases and controls |  |
|  |  | Cross-sectional study - give the eligibility criteria, and the sources and methods of selection of participants | The study included individuals who had a verified history of stroke.  Participants were recruited from individuals utilizing the Holter service.  Participants were categorized into groups depending on whether they had a history of stroke and whether significant findings were detected in the Holter analysis. Those without specific ECG findings were considered as the control group. |
|  |  | Cohort study - for matched studies, give matching criteria and number of exposed and unexposed Case-control study - for matched studies, give matching criteria and the number of controls per case |  |
| Variables | 7 | Clearly define all outcomes, exposures, predictors, potential confounders, and effect modifiers; give diagnostic criteria, if applicable | The study focused on identifying significant findings in the 24-hour three-channel Holter ECG results of post-stroke patients as the primary outcomes of interest. The exposure of interest was the history of stroke among the participants.  Age, gender, symptoms, test duration and duration post stroke were examined as predictors and effect modifiers to assess their impact on the detection rate of significant ECG findings.  Inconsistencies in symptom reporting were considered potential confounders in the study, as highlighted in the limitation section. The diagnostic criteria relied on the presence of significant Holter ECG findings. |
| Data sources/measurement | 8* | For each variable of interest, give sources of data and details of methods of assessment (measurement); describe comparability of assessment methods if there is more than one group | The primary data source for identifying significant findings was the ECG recordings.  Following the exclusion of recordings with artefacts, the recorded data were analyzed using Cardioscan 12 satellite software. Trained cardio physiologists then reviewed the data, categorizing post-stroke patients into groups based on the presence or absence of significant findings.  Uniform methods and protocols were applied to assess all participants, ensuring consistency across the study population. |
| Bias | 9 | Describe any efforts to address potential sources of bias | The exclusion of recordings with artefacts helped mitigate potential bias arising from technical issues. The review of data by trained cardio physiologists helped minimize bias introduced by inexperienced assessors. |
| Study size | 10 | Explain how the study size was arrived at | The study encompassed all available Holter results of post-stroke subjects globally within the institution. |
| Quantitative variables | 11 | Explain how quantitative variables were handled in the analyses; if applicable, describe which groupings were chosen and why | Age data were presented as median and interquartile range, and Mann-Whitney tests were employed to compare age differences between groups with and without significant findings. This approach and statistical test were chosen due to the non-normal distribution of the data.  Participants were divided into groups based on the presence or absence of significant ECG findings, facilitating comparisons between these groups. Additionally, subgroup analyses were conducted based on age, gender, and symptoms to explore the influence of these factors on the detection rate of significant ECG findings. |
| Statistical methods | 12 | Describe all statistical methods, including those used to control for confounding | Statistical methods included normality testing, descriptive statistics using median and interquartile range, Mann-Whitney tests, chi-square tests, and multivariate logistic regression. These methods were selected to address the non-normal distribution of the data and to control for potential confounding variables, thereby enhancing the robustness of the analysis. |
|  |  | Describe any methods used to examine subgroups and interactions | Subgrouping based on age group, gender, and symptoms was conducted to explore their influence on the detection rate of significant ECG findings. Multivariate logistic regression was utilized to examine the relationship between the predictor variables. |
|  |  | Explain how missing data were addressed | Limitations were noted regarding potential inconsistencies in symptom reporting, which could result in missing data. Readers were cautioned to consider these limitations when generalizing the study findings.  Records lacking pertinent information, such as age and gender, or the stroke dates outside the specified duration were excluded from the analysis to ensure data reliability and accuracy. |
|  |  | Cohort study - if applicable, explain how loss to follow-up was addressed |  |
|  |  | Case-control study - if applicable, explain how matching of cases and controls was addressed |  |
|  |  | Cross-sectional study - if applicable, describe analytical methods taking account of sampling strategy | The analytical method was previously described in item #12. Convenient sampling was utilized, encompassing all available Holter results of post-stroke subjects worldwide within the institution. For the control group, stratified sampling was employed to select subjects matched for gender and age with the patients. |
|  |  | Describe any sensitivity analyses | Sensitivity analyses were not conducted as part of this study. |
| Results |  |  |  |
| Participants | 13* | Report numbers of individuals at each stage of study - e.g., numbers potentially eligible, examined for eligibility, confirmed eligible, included in the study, completing follow-up, and analyzed | A total of 6,411 records were initially screened for eligibility, resulting in 978 records meeting the eligibility criteria and being included in the analysis. No follow-up was conducted given the cross-sectional nature of the study. |
|  |  | Give reasons for nonparticipation at each stage Consider use of a flow diagram | A flow chart (Figure 1) illustrating the study's progression was provided in the manuscript. |
| Descriptive data | 14* | Give characteristics of study participants (e.g., demographic, clinical, social) and information on exposures and potential confounders | In the process of record selection, a total of 98 patients and 98 controls were enrolled for the first and second parts of the study, respectively. Table 1 presents the median age and gender distribution. For the third part of the study, 779 patients were included, with a median age of 67 years and an interquartile range of 20 years. Among these participants, 438 were male and 347 were female.  Regarding clinical characteristics, Table 1 provides counts and percentages of significant findings between stroke patients and controls. Table 2 displays various significant findings among stroke patients, along with their age, gender, and symptoms.  Exposures were depicted in Figure 2, showing the week-specific distribution of individuals with significant ECG findings following a stroke. Figure 3 illustrated the month-specific distribution of individuals with significant ECG findings following a stroke, undergoing Holter testing with different durations.  In terms of potential confounders, factors such as inconsistencies in symptom reporting were identified as potential confounders, as discussed in the limitation section. |
|  |  | Indicate number of participants with missing data for each variable of interest Cohort study - summarize follow-up time (e.g., average and total amount) | Records lacking relevant information, including age and gender (35 records), were excluded from the analysis. Additionally, records without recorded stroke dates were also excluded.  Since the study followed a cross-sectional design, there was no follow-up period. |
| Outcome data | 15* | Cohort study - report numbers of outcome events or summary measures over time |  |
|  |  | Case-control study - report numbers in each exposure category, or summary measures of exposure |  |
|  |  | Cross-sectional study - report numbers of outcome events or summary measures | Table 1 displays the frequency of outcome events or summary measures, representing the number of individuals with different significant findings among stroke patients and controls.  Table 2 exhibits the frequency of individuals with various significant findings, accompanied by their median age, male proportion, and the percentage of subjects experiencing symptoms, notably palpitations, syncope, and dizziness.  Appendices II and III in the supplementary file contain the event data and totals of various significant findings, which are utilized to construct Figures 2 and 3. |
| Main results | 16 | Give unadjusted estimates and, if applicable, confounder-adjusted estimates and their precision (e.g., 95% confidence interval); make clear which confounders were adjusted for and why they were included Report category boundaries when continuous variables were categorized | Unadjusted estimates of age, male proportion, and the percentage of individuals experiencing symptoms related to various significant ECG findings were provided.  Adjusted estimates were derived through multivariate logistic regression analyses, investigating the odds ratios of age, gender, and symptoms towards significant ECG findings, along with their respective 95% confidence intervals. These variables were selected to account for potential confounding effects among each other.  Additionally, multiple linear regression analyses were conducted to assess the 95% confidence intervals of the estimates of Holter monitoring duration and duration since stroke onset towards the detection rate of significant ECG findings. |
|  |  | If relevant, consider translating estimates of relative risk into absolute risk for a meaningful time period | The results were not only presented as estimates of relative risk but also in absolute risk, as demonstrated in the interpretation of results in Figure 2. The interpretation of the findings encompassed correlation analysis alongside specific detection rates of certain significant findings. |
| Other analyses | 17 | Report other analyses done - e.g., analyses of subgroups and interactions, and sensitivity analyses | Subgrouping based on age group, gender, and symptoms was conducted to explore the influence of these factors on the detection rate of significant ECG findings. Multivariate logistic regression and multiple linear regression facilitated the examination of the relationship between the predictor variables.  No sensitivity analyses were conducted in this study. |
| Discussion |  |  |  |
| Key results | 18 | Summarize key results with reference to study objectives | In the discussion, the main findings are synthesized in accordance with the study's aims, which include comparing AF prevalence between stroke patients and controls, identifying factors associated with arrhythmias, investigating temporal patterns of cardiac arrhythmias post-stroke, and assessing the effectiveness of extended monitoring for AF and PF detection. |
| Limitations | 19 | Discuss limitations of the study, taking into account sources of potential bias or imprecision; discuss both direction and magnitude of any potential bias | The discussion section critically evaluates the limitations of the research, notably highlighting the potential for inconsistency in symptom reporting by referring physicians and the variability in treatment received by patients as sources of bias. Specifically, the discussion underscores how inconsistent symptom reporting may lead to an underestimation of symptom prevalence among stroke patients. |
| Interpretation | 20 | Give a cautious overall interpretation of results considering objectives, limitations, multiplicity of analyses, results from similar studies, and other relevant evidence | The discussion addresses the objectives by highlighting the elevated detection rate of atrial fibrillation (AF) in stroke patients compared to controls, consistent with prior research. Age and symptomatology emerge as significant factors influencing AF prevalence among stroke patients, with older individuals and those presenting symptoms showing higher detection rates. Additionally, limitations are acknowledged (see item #19), cautioning against broad generalizations of the findings. The study also contextualizes its results by referencing similar studies and reinforcing its findings with corroborative evidence. For instance, regarding the impact of age on detection rates, the study cites research by Kamel et al., which reported a decrease in hazard ratio of AF with age. |
| Generalizability | 21 | Discuss the generalizability (external validity) of the study results | The study findings possess a degree of generalizability since they encompass data from multiple countries and include patients across various stroke severities and age ranges. However, the study also acknowledges limitations that may impact the broader applicability of the results (refer to item #19). |
| Other information |  |  |  |
| Funding | 22 | Give the source of funding and the role of the funders for the present study and, if applicable, for the original study on which the present article is based | Funding for this study was provided by Cardioscan Pye Ltd. The role of the funder was solely in financial support and did not extend to study design, data collection, analysis, interpretation, or manuscript preparation. |

Appendix II

95% confidence interval of median of table 1 in research paper

|  | N | Age, confidence interval |
| --- | --- | --- |
| Total significant finding | 26 | 69, 75 |
| without significant finding | 72 | 52, 69 |
| with AF or PF | 23 | 69, 75 |
| without AF or PF | 75 | 52, 70 |
| with AF | 20 | 69, 78 |
| without AF | 78 | 53, 70 |
| with PF | 3 | 70, 75 |
| without PF | 95 | 59, 71 |
| with SV | 1 | 75, 75 |
| without SV | 97 | 60, 71 |
| with VT | 2 | 66, 80 |
| without VT | 96 | 59, 71 |

Appendix II

| Week since stroke | Total | AF | | CB | | PA | | PF | | SD | |
| --- | --- | --- | --- | --- | --- | --- | --- | --- | --- | --- | --- |
|  |  | Event | % | Event | % | Event | % | Event | % | Event | % |
| 1 | 217 | 25 | 11.5 | 1 | 0.5 | 5 | 2.3 | 4 | 1.8 | 1 | 0.5 |
| 2 | 154 | 5 | 3.2 | 1 | 0.6 | 2 | 1.3 | 5 | 3.2 | 1 | 0.6 |
| 3 | 204 | 1 | 0.5 | 0 | 0 | 0 | 0 | 7 | 3.4 | 2 | 1 |
| 4 | 88 | 0 | 0 | 0 | 0 | 2 | 2.3 | 1 | 1.1 | 1 | 1.1 |
| 5 | 60 | 1 | 1.7 | 0 | 0 | 0 | 0 | 2 | 3.3 | 0 | 0 |
| 6 | 54 | 1 | 1.9 | 0 | 0 | 0 | 0 | 1 | 1.9 | 1 | 1.9 |
| 7 | 48 | 0 | 0 | 0 | 0 | 1 | 2.1 | 1 | 2.1 | 0 | 0 |
| 8 | 32 | 0 | 0 | 0 | 0 | 0 | 0 | 0 | 0 | 0 | 0 |
| 9 | 21 | 0 | 0 | 0 | 0 | 1 | 4.8 | 3 | 14.3 | 1 | 4.8 |
| 10 | 12 | 0 | 0 | 0 | 0 | 0 | 0 | 2 | 16.7 | 0 | 0 |
| Correlation coefficient |  |  | -0.769 |  | -0.683 |  | -0.169 |  | 0.333 |  | -0.169 |
| p-value |  |  | 0.009 |  | 0.029 |  | 0.641 |  | 0.349 |  | 0.641 |

Cont.

| Week since stroke | Total | SV | SV | VE | VE | VT | VT | Total significant findings | Total significant findings |
| --- | --- | --- | --- | --- | --- | --- | --- | --- | --- |
| 1 | 217 | 2 | 0.9 | 0 | 0 | 6 | 2.8 | 44 | 20.3 |
| 2 | 154 | 2 | 1.3 | 0 | 0 | 3 | 1.9 | 19 | 12.3 |
| 3 | 204 | 7 | 3.4 | 1 | 0.5 | 8 | 3.9 | 26 | 12.7 |
| 4 | 88 | 0 | 0 | 2 | 2.3 | 3 | 3.4 | 9 | 10.2 |
| 5 | 60 | 1 | 1.7 | 0 | 0 | 6 | 10 | 10 | 16.7 |
| 6 | 54 | 2 | 3.7 | 0 | 0 | 2 | 3.7 | 7 | 13 |
| 7 | 48 | 2 | 4.2 | 2 | 4.2 | 5 | 10.4 | 11 | 22.9 |
| 8 | 32 | 3 | 9.4 | 0 | 0 | 2 | 6.3 | 5 | 15.6 |
| 9 | 21 | 0 | 0 | 0 | 0 | 2 | 9.5 | 7 | 33.3 |
| 10 | 12 | 0 | 0 | 0 | 0 | 1 | 8.3 | 3 | 25 |
| Correlation coefficient |  |  | -0.006 |  | -0.127 |  | 0.697 |  | 0.624 |
| p-value |  |  | 0.987 |  | 0.727 |  | 0.031 |  | 0.06 |

Appendix III

|  |  |  | AF | | PF | | other significant findings | |
| --- | --- | --- | --- | --- | --- | --- | --- | --- |
| Duration of Holter testing (days) | Duration since stroke onset (months) | Total | Event | % | Event | % | Event | % |
| 1 | 1 | 100 |  | 0.00 |  | 0.00 | 7 | 0.07 |
| 1 | 2 | 42 |  | 0.00 |  | 0.00 | 2 | 0.05 |
| 1 | 2.5 | 1 |  | 0.00 |  | 0.00 |  | 0.00 |
| 2 | 1 | 23 | 2 | 0.09 |  | 0.00 |  | 0.00 |
| 2 | 2 | 7 |  | 0.00 | 1 | 0.14 | 3 | 0.43 |
| 3 | 1 | 111 | 1 | 0.01 | 3 | 0.03 | 9 | 0.08 |
| 3 | 2 | 18 |  | 0.00 | 1 | 0.06 | 1 | 0.06 |
| 3 | 2.5 | 4 |  | 0.00 | 1 | 0.25 |  | 0.00 |
| 4 | 1 | 14 |  | 0.00 |  | 0.00 | 1 | 0.07 |
| 4 | 2 | 3 |  | 0.00 |  | 0.00 | 1 | 0.33 |
| 5 | 1 | 18 |  | 0.00 |  | 0.00 | 1 | 0.06 |
| 5 | 2 | 6 |  | 0.00 |  | 0.00 | 1 | 0.17 |
| 6 | 1 | 22 | 1 | 0.05 | 1 | 0.05 |  | 0.00 |
| 6 | 2 | 5 |  | 0.00 |  | 0.00 | 1 | 0.20 |
| 6 | 2.5 | 2 |  | 0.00 |  | 0.00 |  | 0.00 |
| 7 | 1 | 224 | 5 | 0.02 | 6 | 0.03 | 23 | 0.10 |
| 7 | 2 | 46 | 1 | 0.02 | 1 | 0.02 | 5 | 0.11 |
| 7 | 2.5 | 4 |  | 0.00 |  | 0.00 | 1 | 0.25 |
| 8 | 1 | 3 |  | 0.00 |  | 0.00 |  | 0.00 |
| 8 | 2 | 1 |  | 0.00 |  | 0.00 |  | 0.00 |
| 9 | 1 | 6 |  | 0.00 | 1 | 0.17 |  | 0.00 |
| 9 | 2 | 4 |  | 0.00 |  | 0.00 |  | 0.00 |
| 10 | 1 | 3 |  | 0.00 | 1 | 0.33 |  | 0.00 |
| 10 | 2 | 1 |  | 0.00 |  | 0.00 |  | 0.00 |
| 11 | 2.5 | 1 |  | 0.00 |  | 0.00 | 1 | 1.00 |
| 11 | 1 | 7 |  | 0.00 | 1 | 0.14 |  | 0.00 |
| 12 | 1 | 10 |  | 0.00 | 1 | 0.10 | 3 | 0.30 |
| 12 | 2 | 6 |  | 0.00 | 1 | 0.17 | 1 | 0.17 |
| 13 | 1 | 8 |  | 0.00 |  | 0.00 | 2 | 0.25 |
| 13 | 2 | 7 |  | 0.00 |  | 0.00 | 2 | 0.29 |
| 13 | 2.5 | 1 |  | 0.00 |  | 0.00 |  | 0.00 |
| 14 | 1 | 21 |  | 0.00 |  | 0.00 | 2 | 0.10 |
| 14 | 2 | 48 | 1 | 0.02 | 1 | 0.02 | 10 | 0.21 |
| 14 | 2.5 | 7 |  | 0.00 | 1 | 0.14 | 2 | 0.29 |
|  | Grand Total | 779 | 11 | 0 | 21 | 0.03 |  | 0 |

Appendix IV

Combined effect size of atrial fibrillation or paroxysmal atrial fibrillation prevalence, 24-72 hours post-stroke in prior studies

| Study | Event | Total | Prevalence | Standard error* | Number |
| --- | --- | --- | --- | --- | --- |
| Our study | 23 | 98 | 0.23 | 0.05 | 98 |
| (Wohlfahrt et al., 2014) | 42 | 224 | 0.19 | 0.03 | 224 |
| (Fujii et al., 2013) | 45 | 288 | 0.15 | 0.02 | 215 |
| (Sutamnartpong et al., 2014) | 139 | 1240 | 0.24 | 0.03 | 204 |
| (Yoshioka et al., 2015) | 48 | 204 | 0.16 | 0.03 | 288 |
| (Giralt-Steinhauer et al., 2015) | 174 | 2490 | 0.11 | 0.01 | 1240 |
| (Kamel et al., 2009) | 32 | 215 | 0.07 | 0.03 | 2490 |
| (Amaya Pascasio et al., 2022) | 102 | 460 | 0.22 | 0.02 | 460 |
| Combined effect size (excluding our study) |  |  | 0.16 | 0.02 | 5219 |

* Standard error was computed using the formula: SQRT(total)/event.
